# Community factors associated with local RSV epidemic patterns: a spatiotemporal modeling study

**DOI:** 10.1101/2020.07.06.20144345

**Authors:** Zhe Zheng, Virginia E. Pitzer, Joshua L. Warren, Daniel M. Weinberger

## Abstract

**Background:** Respiratory syncytial virus (RSV) causes a large burden of morbidity in infants, young children, and the elderly. The timing of RSV seasonal epidemics exhibits strong spatial patterns within the United States. Spatial variability in the timing of RSV epidemics provides an opportunity to probe the factors driving transmission of the virus.

**Methods:** We evaluated competing hypotheses about the associations between RSV epidemic timing at the ZIP-code level and household size, population density, school district boundaries, commuting patterns, and geographic proximity. We used hierarchical Bayesian models with monthly ZIP-code level hospitalization data from New York, New Jersey, and Connecticut between July 1997 and June 2014 to investigate these hypotheses.

**Results:** Early epidemic timing across ZIP codes was associated with large household sizes and high population density, and nearby ZIP codes shared similar epidemic timing. Variations in epidemic timing attributed to commuting patterns or school district boundaries are negligible.

**Conclusion:** Our results suggest RSV epidemics take off faster in areas with more household crowding and higher population density, and that epidemic spread follows a spatial diffusion process based on geographic proximity. With several vaccines against RSV under development, it is important to understand the drivers of RSV transmission and disease in order to maximize population protection of a vaccine program. Our findings can inform more effective control measures against RSV, such as vaccine programs and household infection control, and guide future studies on the transmission dynamics of RSV.

## INTRODUCTION

Respiratory syncytial virus (RSV) causes over 2 million outpatient visits annually among children in the United States (US)(*1*). It is one of the leading causes of hospitalization for lower respiratory infection among young children and the elderly (*2, 3*). Developing an effective vaccine against RSV is a high priority, as prophylaxis against RSV is prohibitively expensive and complicated to administer(*4-6*). Understanding the community factors associated with the transmission dynamics of RSV can help to design efficient clinical trials for efficacy and effectiveness evaluation and to optimize the implementation of an RSV vaccine program (*7*).

The timing of RSV epidemics varies markedly over space, both over large geographic scales and within a region (*8, 9*). Across the US, epidemics start earliest in the southeast and later to the north and west (*10*). At smaller scales, epidemics tend to start earlier and last longer in urban areas than in surrounding suburbs and rural areas (*6*). Environmental and climatic drivers can explain some of the broad spatial differences in the timing of RSV epidemics, but cannot fully explain finer-scale spatial variations(*11, 12*). Other than environmental and climatic factors, only a few studies have addressed other putative drivers of RSV spatiotemporal dynamics, mostly demographic factors (*9, 13, 14*). A better understanding of the drivers of local RSV spatiotemporal dynamics is needed. Hierarchical Bayesian spatial models offer a flexible platform for accounting for spatial autocorrelation when estimating covariate effects(*15*). We utilize these models to probe the association between the timing of seasonal epidemics of RSV and human mobility, demography, spatial proximity, and school boundaries.

The tri-state area that includes New York, New Jersey, and Connecticut is ideal to study the potential factors driving fine-scale spatial variation in RSV epidemics because of the demographic diversity and high volume of population movement. This area includes New York City, the most populous city in the US. It also includes rural areas like the North Country region of New York state and Litchfield County in Connecticut. In addition, variation in community factors exist at small spatial scales(*16*). The strong commuting ties between New York City and surrounding areas offers valuable opportunities to compare the impacts of commuting flows and geographic distance on RSV transmission.

The principal aim of this study is to determine the relative contributions of demographics, school gathering, geographic proximity, and population movement in RSV transmission. We accomplished this by comparing competing hypotheses characterized by different spatial model structures that build on empirical epidemiologic and sociodemographic data. A better understanding of the drivers of transmission will provide helpful insights in optimizing RSV vaccine administration, informing current disease control guidelines, and planning clinical trials.

## METHODS

### Data source

RSV-specific hospitalization data for children <2 years from 1997 to 2013 were obtained from the Connecticut State Inpatient Database through the Connecticut Department of Public Health (CT-DPH). This dataset included the week and month of admission and the ZIP code of residence. Similar data for New York state and New Jersey were obtained from State Inpatient Databases of the Healthcare Cost and Utilization Project maintained by the Agency for Healthcare Research and Quality; these comprehensive databases contain all hospital discharge records from community hospitals in participating states(*17*). Datasets include the month of hospitalization (for 2005-2014) and ZIP code of residence. Cases were defined as any child <2 years old whose hospital discharge diagnostic codes included 079.6 (RSV), 466.11 (bronchiolitis due to RSV), or 480.1 (pneumonia due to RSV), based on the International Classification of Disease Ninth Revision [ICD-9]. The analysis of the data was approved by the Human Investigation Committees at Yale University and the Connecticut DPH. The authors assume full responsibility for analyses and interpretation of data obtained from the CT-DPH.

Geospatial data at the ZIP-code level were obtained from the US Census Bureau’s Geography program. Information about population size <5 years old and average household size in each ZIP code area were obtained from the US Census Bureau’s American Community Survey. School District information was obtained from state geographic information system and education department databases. Countrywide commuting patterns at the ZIP-code level were obtained from the US Census Bureau’s Center of Economic Studies. All collected demographic and geographic data are from 2010.

### Model structures and hypotheses

#### Two-stage hierarchical Bayesian model

We estimated the phase (peak timing) of RSV epidemics and factors associated with spatial variability in timing at the ZIP-code level using a two-stage modeling approach. In the first stage, we obtained an estimate and corresponding measure of uncertainty of epidemic timing for each ZIP code. In the second stage, we used hierarchical Bayesian spatial models with multiple covariates and different spatial correlation structures to investigate competing hypotheses about the factors that potentially influence epidemic dynamics. Model comparison techniques were used to evaluate the different hypotheses.

The first stage consists of a harmonic Poisson regression model that estimates the amplitude and phase of seasonal RSV epidemics(*18*). This model was fit separately to the monthly time series of observed RSV hospitalizations from each ZIP code and is specified as

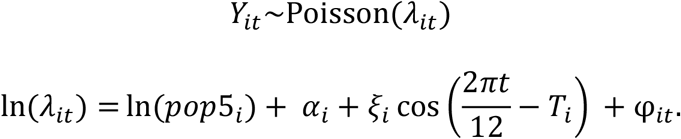

The observed number of RSV cases in ZIP code *i* during month *t* is denoted as *Y*_*it*_, phase *T*_*i*_. captures the shift in peak timing of epidemics (with the 12-month period starting in early July), *ξ*_*i*_ is the amplitude of seasonal epidemics, In (*pop*5_*i*_) is the offset term for the population under 5, *α*_*i*_. is the intercept, and *φ*_*it*_. is an observation-level random effect that accounts for any unexplained variability in the data (i.e., overdispersion). In this first stage, all model parameters are ZIP-code specific. This model was fitted in the Bayesian setting using the rjags package with weakly informative prior distributions specified for all model parameters(*19*). Full details are provided in the Supplement.

Based on estimates of *T*_*i*_. (posterior means) and their uncertainty (posterior standard deviations) obtained from the first stage, we use a Bayesian meta-regression approach in the second stage to explain spatial variability in the epidemic peak timing of RSV(*20-23*). Specifically, we first transform the peak timing parameters, whose values are confined to [0, 2π], to have support on the real line (useful for the second stage regression modeling) such that θ_i_ = In 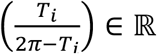. Next, we obtain posterior means,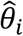, and posterior standard deviations,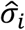, from the first stage and specify the second stage model as 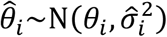 where 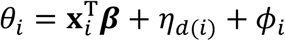

#### Hypotheses

Hypotheses regarding the factors driving the timing of RSV epidemics at a local scale are captured in the second stage model by including covariates (e.g., ZIP-code level household size, population density) and by specifying priors for the random effects that explain the variability. These random effects include school district random effects, independent random effects, and spatially structured random effects that account for correlation defined by spatial neighbors or commuting connectivity. The corresponding model structures for our hypotheses are as follows:

1. There is no spatial correlation in RSV epidemic timing within the three states after adjusting for ZIP-code level covariates and school districts. Timing of RSV epidemics may be affected by ZIP-code average household size and population density, and timing of RSV epidemics is similar for ZIP codes within the same school district, such that *θ*_*i*_ = *β*_*i*_ + *β*_*1*_ ∗ (household size_*i*_) + *β*_2_ ∗ (log pop den_*i*_) + *η*_*d*(*i*)_ + *Φ*_*i*_, where the school district random effects are 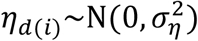 and the nonspatial random effects are 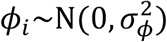. The *d(i)* function maps a ZIP code to its corresponding school district.
2. RSV epidemics have similar timing in neighboring geographic locations and is similar within the same school district, such that *θ*_*i*_ = *β*_0_ + *β*_1_ ∗ (household.size_*i*_) + *β*_2_ ∗ (log pop den_*i*_) + *η*_*d*(*i*)_ + *Φ*_*i*_, where the school district random effects are 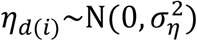 and the spatially structured random effects are 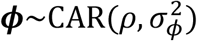(*20*). Here, spatial proximity is defined by ZIP codes with adjacent borders.
3. RSV epidemic timing is explained by commuting flows and is similar within the same school district, such that *θ*_*i*_ = *β*_0_ + *β*_1_ ∗ (household.size_*i*_) + *β*_2_ ∗ (log pop den_*i*_) + *η*_*d*(*i*)_ + *Φ*_*i*_, where the school district random effects are 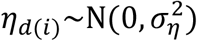 and spatially structured random effects are 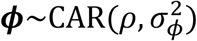(*20*). Here, spatial proximity is defined by commuting patterns across ZIP codes.

More details about hyperprior distributions for these different models can be found in the Supplement.

Parameter estimation was carried out separately for each state via Markov chain Monte Carlo simulation with an initial burn-in period of 10,000 iterations and a subsequent set of 50,000 posterior samples collected for all parameters in the first stage(*24*). A burn-in of 5,000 iterations and a subsequent set of 20,000 posterior samples were collected in the second stage. Convergence was assessed by examining individual parameter trace plots, with no obvious signs of non-convergence observed. Deviance information criterion (DIC) was calculated for each model to compare the performance; a lower DIC indicates a model has an improved balance of fit and complexity(*25*).

We tested different hypotheses regarding variability in peak timing first by testing different spatial correlation structures for *Φ*_*i*_ The spatial structures we evaluated were (1) no spatial correlation, (2) correlation based on geographic proximity, and (3) correlation based on commuting network volume. Then we calculated pseudo R-squared values to measure the role of average household size, population density, spatial connectivity, and school districts in explaining the variation in the data under the spatial structure of the model with the lowest DIC. Finally, we estimated the proportion of residual variability explained by the spatially correlated random effects (*Φ*_*i*_) versus the school district random effects (*η*_*d*(*i*)_) Equations for the pseudo R-squared calculation and attributable percent of residual variability calculation can be found in the Supplement. All analyses were performed using R v3.5.3.

## RESULTS

### Characteristics of the data

In total, we captured 67,244 RSV hospitalizations across 2,612 ZIP codes in New York, New Jersey, and Connecticut. The number of hospitalizations, commuters, and the length of study period varied among states (Table 1). Sociodemographic factors are summarized in Table 2.

**Table 1.** Number of hospitalizations, ZIP codes, school districts, commuters, and study period in New Jersey, New York, and Connecticut.

|  | New Jersey | New York | Connecticut |
| --- | --- | --- | --- |
| RSV Hospitalizations | 19,708 | 38,376 | 9,160 |
| ZIP codes | 592 | 1,745 | 275 |
| School districts* | 337 | 934 | 156 |
| Commuters | 4,295,718 | 8,689,118 | 1,725,973 |
| Study period | July 2005 - June 2014 | July 2005 - June 2014 | July 1997 - June 2013 |

\* We assigned the ZIP codes that do not belong to any school district as a single school district (in total 40) in the analysis.

**Table 2.** Descriptive statistics and sociodemographic factors by ZIP code in New Jersey, New York, and Connecticut, 2010.

|  | Mean (Range) |
| --- | --- |
| Population Size under Five | 713 (1 – 16,177) |
| Population Density (persons/square kilometers)* | 1781.19 (1.69 – 57,248.15) |
| Average Household Size | 3 (2 - 5) |
| Number of Commuters | 14 (2 – 15,598) |

\* log transformed in the analysis

### Spatiotemporal pattern of RSV epidemics

RSV activity began between late fall and early winter in the tri-state area (Figure S1). The epidemics peaked between late winter and early spring. There were variations in RSV epidemic initiation and peak timing between states. The estimated peak timing among ZIP codes ranged from late December (month 5, relative to July) to mid-March (month 8) based on the best fit model (Figure 1). Visually, the local epidemics peaked earliest in urban areas (e.g., the New York metropolitan area) then extended to less populous areas like upper New York state and eastern Connecticut. Epidemic peaks were generally earlier in New Jersey compared to the other states. The overall spatial diffusion trend is similar across models with different assumptions while the assumption of no spatial correlation resulted in a wider range of peak timing estimates (Figure S2).

**Figure 1.**
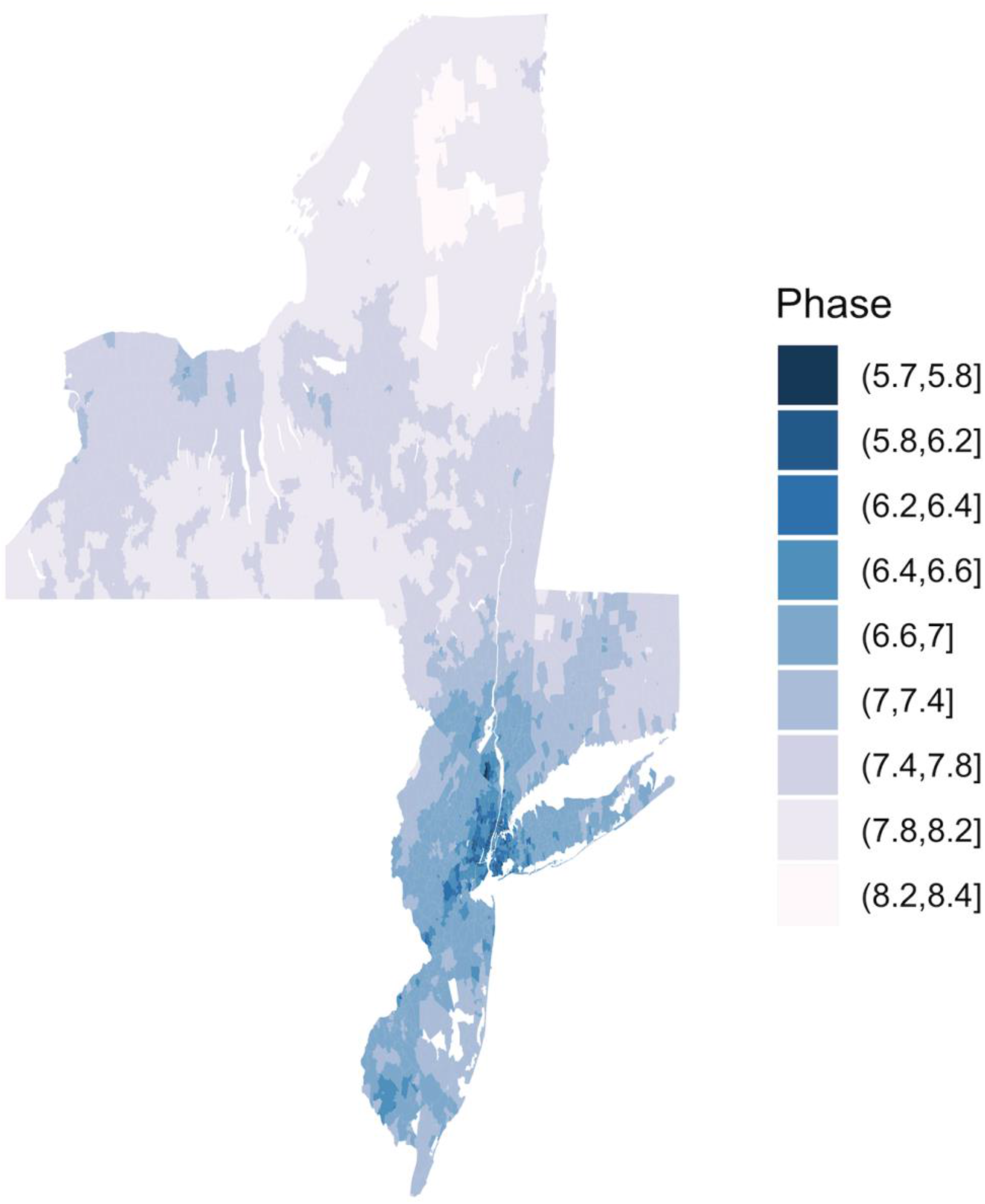
Estimated peak timing of RSV epidemics by ZIP code from the best-fit model. This model accounted for average household size, population density, school district, and geographic proximity. The study periods are July 1997 - June 2013 in Connecticut, July 2005 - June 2014 in New York and New Jersey.

### Major correlates of the spatial variation in RSV epidemics in the three states

Larger average household size and higher population density were associated with earlier epidemic timing (Table S1). Household size and population density explained most of the spatial variability in RSV epidemics timing (Table 3 and Figure S4). These results are consistent between states. In New Jersey, the state with the highest population density in the US, population density is the most important factor in explaining the spatial variability in RSV epidemics, while in Connecticut and New York, average household size ranked first based on the pseudo R-squared analysis (Table 3). The models that accounted for spatial correlation defined by adjacency of neighboring ZIP codes outperformed those defined by commuting flows and those that assumed no spatial correlation (Table 4). More than half of the residual spatial variability in the timing of RSV epidemics was explained by neighboring effects in the tri-state area. In New Jersey and New York, neighboring effects explained 93% and 94% of the residual variability respectively. School district effects contributed little to variations in epidemic timing. (Supplement Table 2). The proportion of variability explained by different factors varied between states (Table 3 and Supplement Table 2).

**Table 3.**
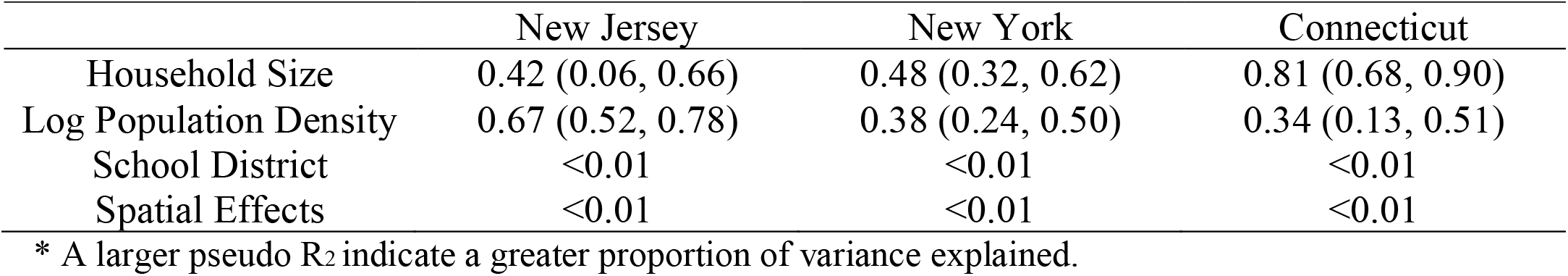
Pseudo R_2_ (Mean and 95% credible interval) of factors associated with spatiotemporal variations in RSV epidemics in New Jersey, New York, and Connecticut*

**Table 4.** DIC scores of competing models in New Jersey, New York, and Connecticut.

|  | New Jersey | New York | Connecticut |
| --- | --- | --- | --- |
| No Spatial Correlation | 1156 | 6454 | 475 |
| Spatial Proximity | 1133 | 6345 | 469 |
| Commuting Flows | 1151 | 6449 | 475 |

## DISCUSSION

With new vaccines against RSV under development, it is critical to understand the transmission patterns of RSV to optimize vaccine deployment. Applying spatial analysis and comparing competing model structures, we set out to better understand the underlying community factors associated with spatial dissemination and timing of RSV epidemics. Our results suggest that epidemics peak earlier in ZIP codes with larger average household size and higher population density, and that epidemics in one location are correlated with those in neighboring areas. Our analysis also shows that RSV epidemics are highly spatially structured and demonstrate considerable variation, beginning in large urban areas and radially spreading to rural places over more than 2months in the tri-state area.

Household size, population density, and geographic proximity were best correlated with RSV spatiotemporal dynamics. Our findings indicate that crowded living conditions might contribute to disease transmission and high population density might precipitate the epidemic outbreaks. These results could be explained by the high contact rates within large households and crowed urban areas. Frequent contacts lead to rapid disease spread and epidemic growth, consistent with a positive correlation between population density and estimated RSV transmission rates for different US states(*11*). In addition, the repeated infection challenges from an epidemic outbreak in a proximate community will eventually form a successful chain of infection and result in the localized and radially diffusive epidemics(*26*). Other factors, such as gathering within schools and commuter flows, might also contribute to RSV transmission, but are not the major drivers of spatiotemporal patterns.

The results from our study have important implications for planning clinical trials and developing transmission dynamic models for RSV. First, since household size may be associated with the risk of exposure and transmission of RSV, vaccine trials should ensure that household size is balanced between vaccinees and non-vaccinees. Second, transmission dynamic models should account for spatial heterogeneity as well as spatial correlations in the force of infection (e.g., using a meta-population model). The spatial patterns of RSV epidemic timing from the best-fit model show considerable discrepancies between urban and rural areas, highlighting the need to consider these as distinct spatial units in RSV transmission models.

Our findings are consistent with previous genomic analyses of RSV, which found that household transmission is common and viruses from nearby households share similar phylogenetic origins(*27, 28*). However, due to the small sample size and study design, genomic analyses were unable to compare the different transmission environments and their relative role in local RSV transmission(*27, 28*). Using empirical epidemiologic, demographic and commuting flow data, our research has found evidence to fill this knowledge gap. Other epidemiological studies also concluded that household crowding and/or a larger number of siblings are associated with increased risk of severe RSV lower respiratory tract infection(*29*). These findings together provide a more complete picture of the major drivers of RSV transmission and how local risk factors affect regional patterns of RSV epidemic spread.

Notably, the spatial spread pattern of RSV epidemics differs from that of influenza(*26, 30, 31*), which suggest the different roles that each age group might play in RSV transmission and influenza transmission(*32*). As a result, the level of indirect protection that might be generated by vaccinating different age groups is expect to vary for the two pathogens(*33, 34*). Spatial studies suggested that commuting flows drive the spread of seasonal influenza epidemics, while school-age children may be the major driver of 2009 H1N1 pandemic in the US(*26, 30, 31*). However, our analysis suggests that short-range modes of transmission, especially household transmission and local community transmission, predominates for RSV spread. Combined with the fact that children under five have the highest relative risk of RSV infection(*32*), our results suggest that infant and young children before school age might play an important role in RSV transmission. This could potentially be explained by the build-up of partial immunity against RSV due to previous exposure as age increases (*35*).

Our study and interpretations have several caveats. First, we performed an ecological analysis with aggregate data on ZIP-code level sociodemographic characteristics. Thus, we did not assess the role of individual household size on risk of RSV transmission, for example. Second, the aggregation of ZIP-code level cases to dominant school districts may have led to some misclassification, and does not account for the multiple primary schools typically found within a school district. This could have affected the data-fitting process and resulted in a lower percentage of variability that was explained by school districts. Third, monthly time series may disguise some detail variations and therefore the underlying mechanism compared to weekly time series. However, in a sensitivity analysis, we estimated the peak epidemic timing in Connecticut using both weekly data and data aggregated to the monthly level, and found that the phase estimates were identical. Thus, the availability of finely temporally-resolved surveillance data is not essential to capture the pattern of disease spread. Fourth, while we draw inference about the major transmission environment and potential mechanism of spread, we did not formally evaluate the relative role of each age group in RSV transmission. The pattern of spatial diffusion that we observed may result from a wide range of possibilities. Thus, the interpretation of our study results needs to be further explored with genomic analyses and transmission dynamic models, as well as epidemiological studies. More detailed data, such as the proportion of young children attending daycare in each ZIP code, could help to address remaining gaps in the knowledge of this system.

In conclusion, our results reveal significant variation in local RSV epidemic timing. We find evidence that the timing of RSV epidemics is associated with average household size, population density, and epidemic timing in neighboring areas. In general, RSV epidemics take off earlier in urban areas and spread to rural places with low population density and average household size. These findings highlight the need for infection control within households and communities to protect high-risk populations. Our results also offer additional insights that can be used to inform the development of transmission dynamic models and provide guidance on future vaccine target populations and clinical trial design.

## SUPPLEMENT

### Supplementary Methods

### Supplementary Results

### Supplemental Methods

#### Two-stage model to estimate epidemic peak timing

##### First Stage

The first stage model is described by the following equations:

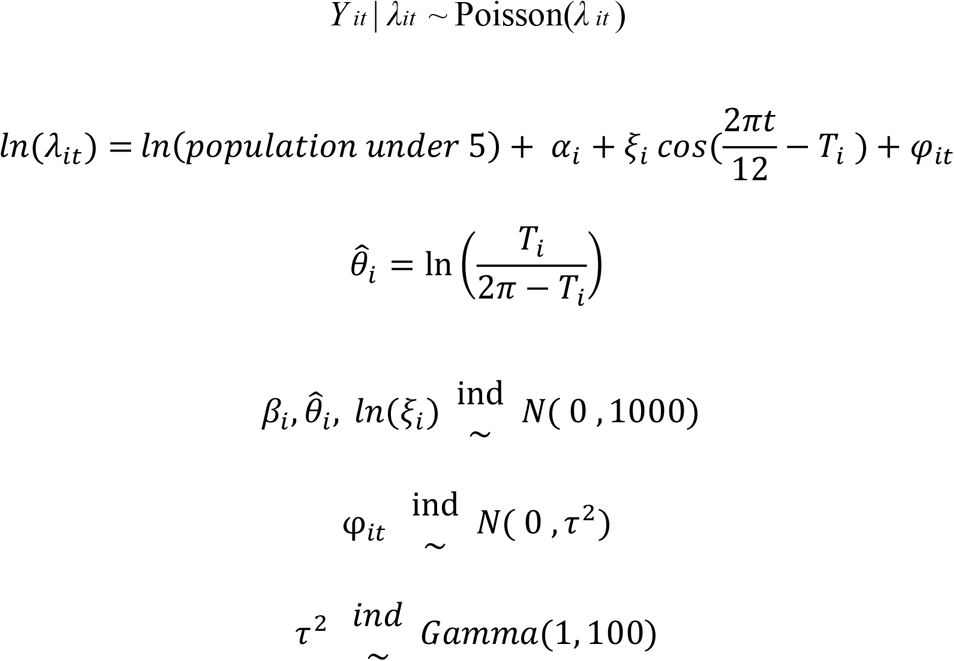

The model variables are defined as:

- *t*: time since July 1997 (in months)
- *n*: number of ZIP code areas
- *q*: number of school districts
- *T*_*i*_: phase or horizontal offset, which represents the epidemic peak timing for ZIP code area *i, T*_*i*_ ∈ [0, 2 π]
- 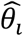: a transform of *T*_*i*_. in order to expand the sampling area to ***R***
- *ξ*_*i*_: the amplitude of seasonal variation
- *β*_*i*_: ZIP-code-level random effect
- *φ*_*it*_: random error (noise) of the series about the period component

## Second Stage

<u>(1) First hypothesis</u>: There is no spatial correlation in RSV epidemic timing within the three states after adjusting for ZIP code-level covariates and shared variability within school districts. In order to obtain desired levels of statistical precision and avoid extreme rates based on small local sample sizes, the non-spatial model uses an *i*.*i*.*d*. random intercept to pool information across all small areas, then the model pulls estimates toward a global mean without regard to their relative spatial locations. This leads to the following formulation for the estimate of the epidemic peak timing, *θ*_*i*_:

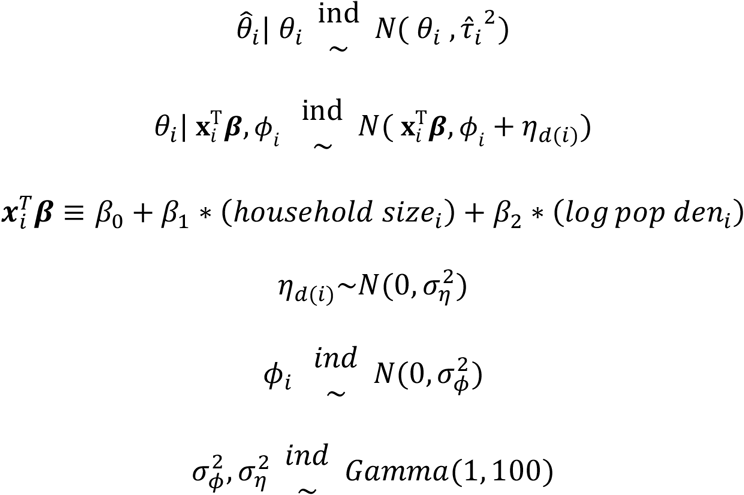

The model variables are defined as:

- 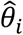: the phase estimates of each ZIP code area from the first stage
- 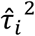: the variance of 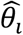, which can be calculated from the posterior samples of the first stage model.
- *Φ*_*i*_: ZIP-code-level random effect
- 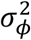: variance of ZIP code random effect
- *η*_*d*(*i*)_: school district random effect
- 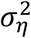: variance of school district random effect

<u>(2) Second hypothesis</u>: RSV epidemics have similar timing in neighboring geographic locations after adjusting for ZIP code-level covariates and shared variability within the same school district. In this hypothesis, we adopted a conditional autoregressive (CAR) prior model with a Leroux formulation for smoothing the spatial random effect term, which allowed the models to have a single random intercept accounting for both spatially structured and unstructured variation(*20*). The degree of smoothing in spatial and non-spatial components of the random effect was determined by the empirical data. Spatial correlation was estimated by *ρ*. The neighborhood matrix (*W*\*) contained information about spatial proximity between two locations. Here, spatial proximity is defined as ZIP codes with adjacent borders.

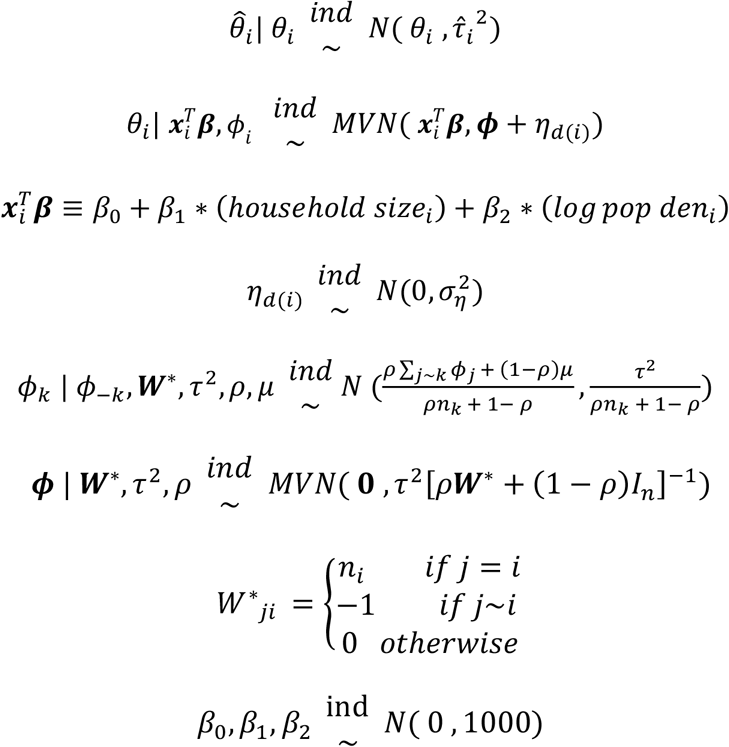

- 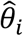: the phase estimates of each ZIP code area from the first stage
- 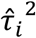: the variance of 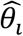, which can be calculated from the posterior samples of the first stage model.
- *θ*_*i*_: ZIP code level true epidemic peak timing
- *n*_*i*_: the number of ZIP code areas that share the border with ZIP code area *i*
- ***W***^**∗**^: an n ⨯ n neighborhood matrix. The diagonal corresponds to how many neighboring ZIP codes that ZIP code *i* has. If ZIP code areas *i* and *j* share a border,*W*\*_*ji*_ = −1, otherwise *W*\*_*ji*_ = 0.
- *I*_*n*_ Identity matrix
- *ρ*: spatial autocorrelation parameter; *ρ* ∈ [0, 1), where *ρ* = 0 corresponds to spatial independence, simplifying the model to an independent random effects model, and *ρ* near 1 means strong spatial autocorrelation. However, *ρ* should not equal to 1, since this corresponds to an improper intrinsic autoregressive (IAR) model(*21, 36*).
- *τ*^2^: variance of ZIP-code-level random effect
- *η*_*d*(*i*)_: school district random effect
- 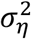: variance of school district random effect

<u>(3) Third hypothesis</u>: RSV epidemic timing is explained by commuting flows after adjusting for ZIP code-level covariates and school districts. The model structure is similar to the second hypothesis, but instead of defining the neighborhood matrix based on shared geographic borders, spatial proximity in the commuting model (***W***) is defined by the frequency of commuters between two locations.

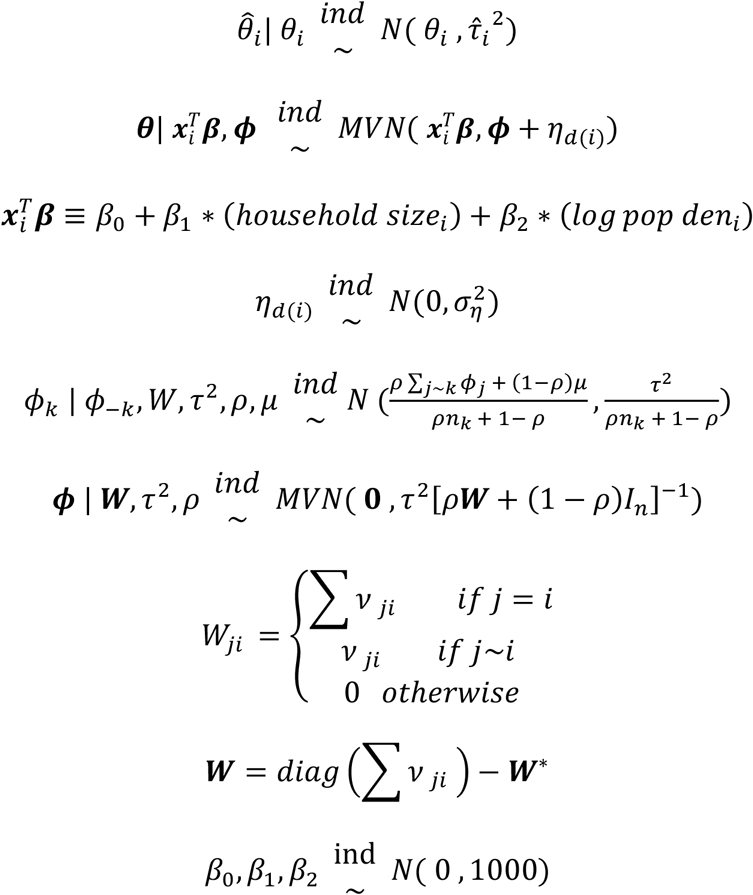

- 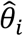: the phase estimates of each ZIP code area from the first stage
- 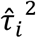: the variance of 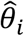 which can be calculated from the posterior samples of the first stage model.
- *θ*_*i*_: ZIP code level true epidemic peak timing
- *η*_*d*(*i*)_: school district random effect
- 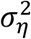: variance of school district effect
- *μ*: an overall mean
- ***W***^**∗**^: an *n* ⨯ *n* neighboring matrix, where the element *v* _*ji*_. is defined by the commuting frequency between ZIP codes *j* and *i*.
- *I*_*n*_: Identity matrix
- *ρ*: spatial autocorrelation parameter; *ρ* ∈ [0, 1)
- *τ*^2^: variance of ZIP-code-level random effect

*Percentage of variability explained by school district:*

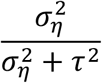

*Percentage of variability explained by spatial correlation:*

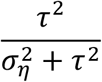

*Pseudo R-square(R*_*2*_*) calculation:*

Reference model Sum of Squares Due to Error (SSE): 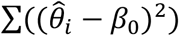

Household size predictor Pseudo *R*_*2*_ Value: 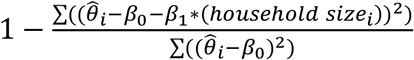

Log population density predictor Pseudo *R*_*2*_ Value: 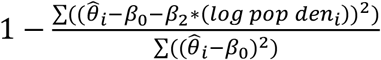

School district random effect Pseudo *R*_*2*_ Value: 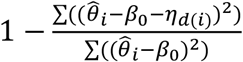

Household size predictor Pseudo *R*_*2*_ Value: 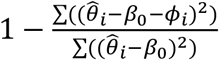

### Sensitivity Analysis

To assure the robustness of our analysis, we compared the phase estimates with monthly aggregated data to the phase estimates with weekly data in Connecticut. We aggregated the original weekly time series data in Connecticut to monthly data based on the month that the first dates of the week fell into. We then ran the two-stage model twice with the two types of dataset. At the end, we measured their similarity by constructing a linear regression and calculating the slope and intercept (Figure S5).

### School District Interpolation

RSV hospitalization cases were reported at ZIP code level. We applied areal interpolation process to derive estimates at school district level from the ZIP code level report. First, we calculated the intersection between ZIP codes and school districts. Then we measured their areal weight and multiplied the value by the weight to get a hospitalization cases estimate for each intersection. At the end, we aggregated these estimated value to get an estimate for each school district(*37*). The analysis was performed using areal package version 0.1.6(*38*).

## Data Availability

The demographic and geographic data that support the findings of this study are available in the Geography program of the US Census Bureau, the American Community Survey of the US Census Bureau, the Center of Economic Studies of the US Census Bureau, state geographic information system, and education department databases. The hospitalization data are not available publicly but can be attained from the State Inpatient Database with the permission of the Connecticut Department of Public Health (CT-DPH) and upon signing a data use agreement with the Agency for Healthcare Research and Quality.

https://www2.census.gov/geo/tiger/TIGER2010/ZCTA5/2010/

https://data.census.gov/cedsci/

https://njogis-newjersey.opendata.arcgis.com/datasets/ca144194df66491d83b8f8bf338e0172_2

https://gis.ny.gov/gisdata/inventories/details.cfm?DSID=1326

http://magic.lib.uconn.edu/connecticut_data.html

https://lehd.ces.census.gov/data/

https://github.com/weinbergerlab/RSV_spatiotemporal

## Supplemental Results of the two-stage hierarchical model

**Figure S1.**
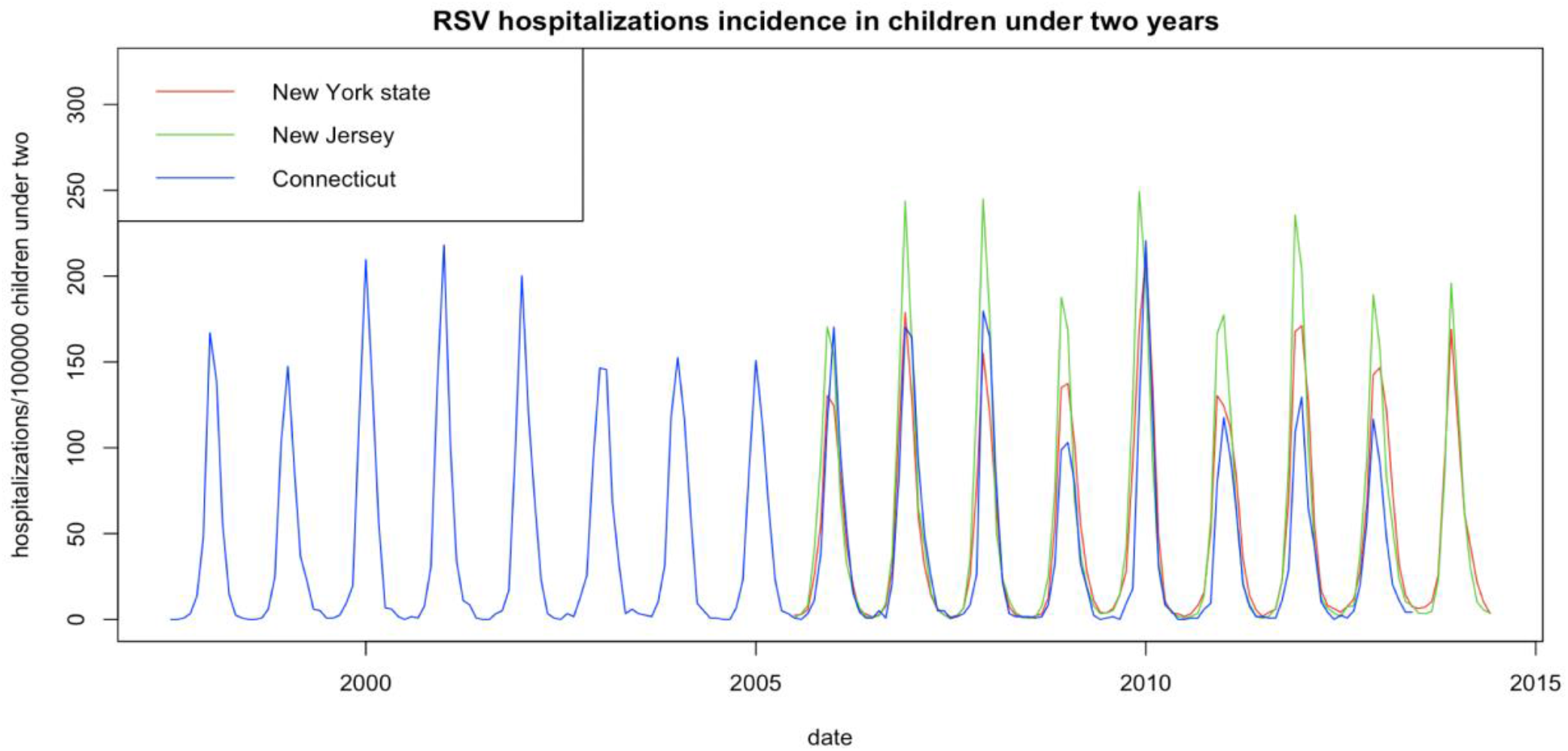
RSV hospitalization incidence in children under two.

**Figure S2.**
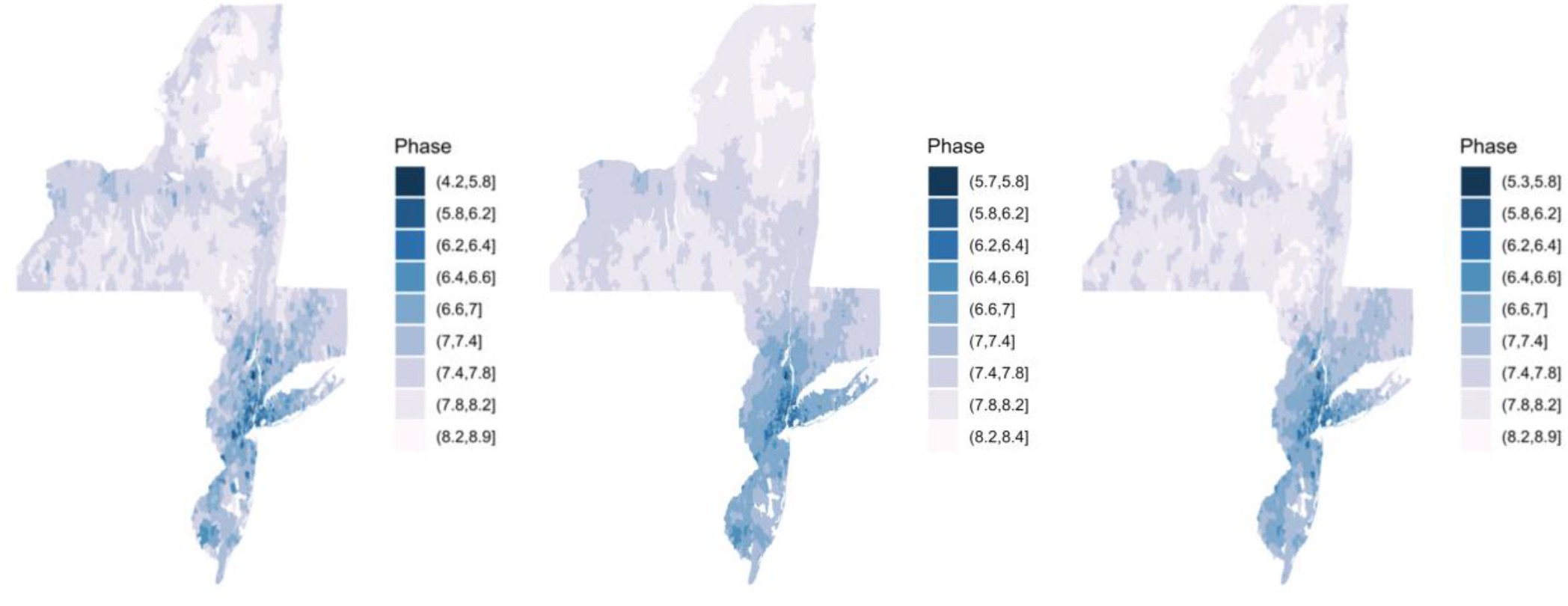
Estimated peak timing of RSV epidemics by ZIP code from the competing models. (A) Model assumed no spatial correlation while accounting for average household size, population density, and school district. (B) The best-fit model. This model accounted for average household size, population density, school district, and geographic proximity (C) Commuting network model. This model accounted for average household size, population density, school district, and commuting flows. The study periods are July 1997 - June 2013 in Connecticut, July 2005 - June 2014 in New York and New Jersey.

**Figure S3.**
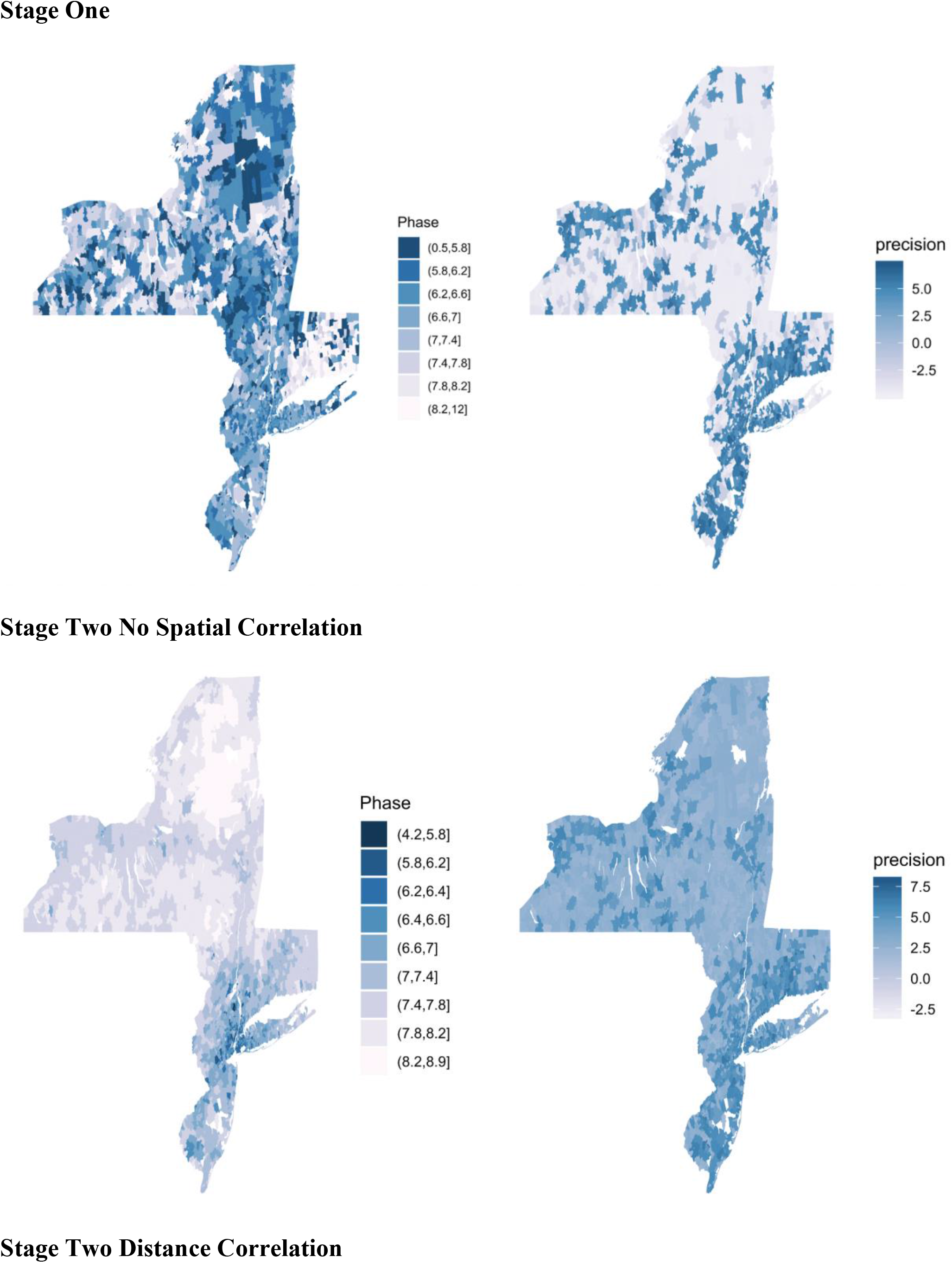
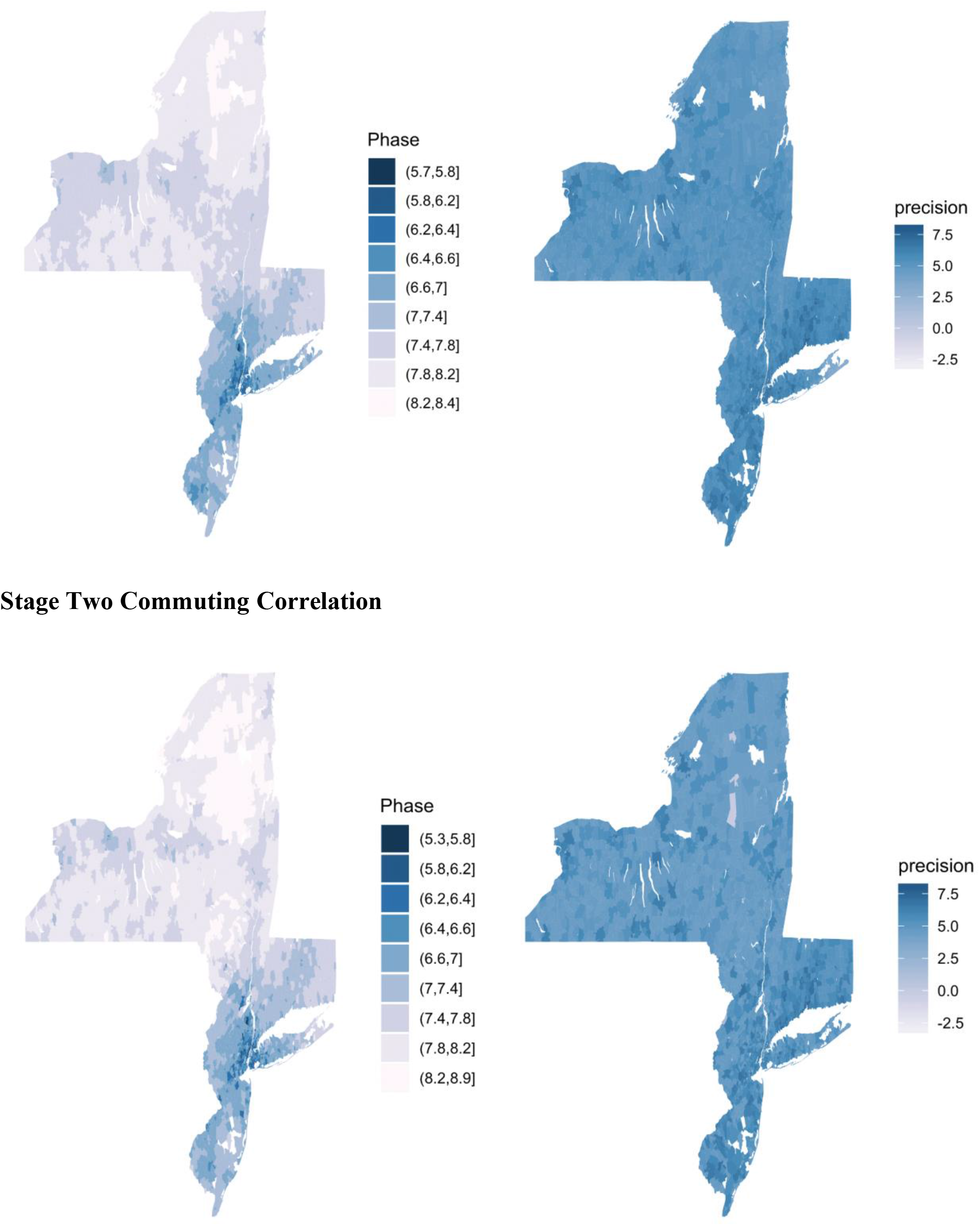
Epidemic Peak Timing Estimation and Precision of Competing Models. The left panel showed the RSV epidemic peak timing estimates by different models. The right panel showed the precision of these estimates. The darker the blue is, the more precise the estimates are. Before applying the second stage model, estimates of peak timing from the first stage model were more variable overall, including a few extreme values. The estimates of many ZIP codes had low precision in the stage 1 model. The geographic distance based spatial correlation structure pool information across nearby small areas, obtaining desired levels of statistical precision and avoid extreme values based on small local sample sizes.

**Supplement Table 1.** Posterior estimation of fixed effects coefficient and spatial correlation coefficient.

|  | New Jersey |  | New York |  | Connecticut |  |
| --- | --- | --- | --- | --- | --- | --- |
|  | Mean | 95% CrI | Mean | 95% CrI | Mean | 95% CrI |
| Household Size | -0.08 | (-0.13, -0.03) | -0.09 | (-0.12, -0.05) | -0.26 | (-0.35, -0.18) |
| Population Density | -0.06 | (-0.05, -0.03) | -0.03 | (-0.04, -0.02) | -0.03 | (-0.05, -0.01) |
| $\rho^*$ | 0.94 | (0.75, 1.00) | 0.99 | (0.99, 1.00) | 0.80 | (0.30, 0.99) |

The posterior mean of the spatial autocorrelation parameter, $\rho$ , was $\geq 0.8$ across the three states, suggesting that the residual variability was spatially structured.

**Supplemental Figure 4.**
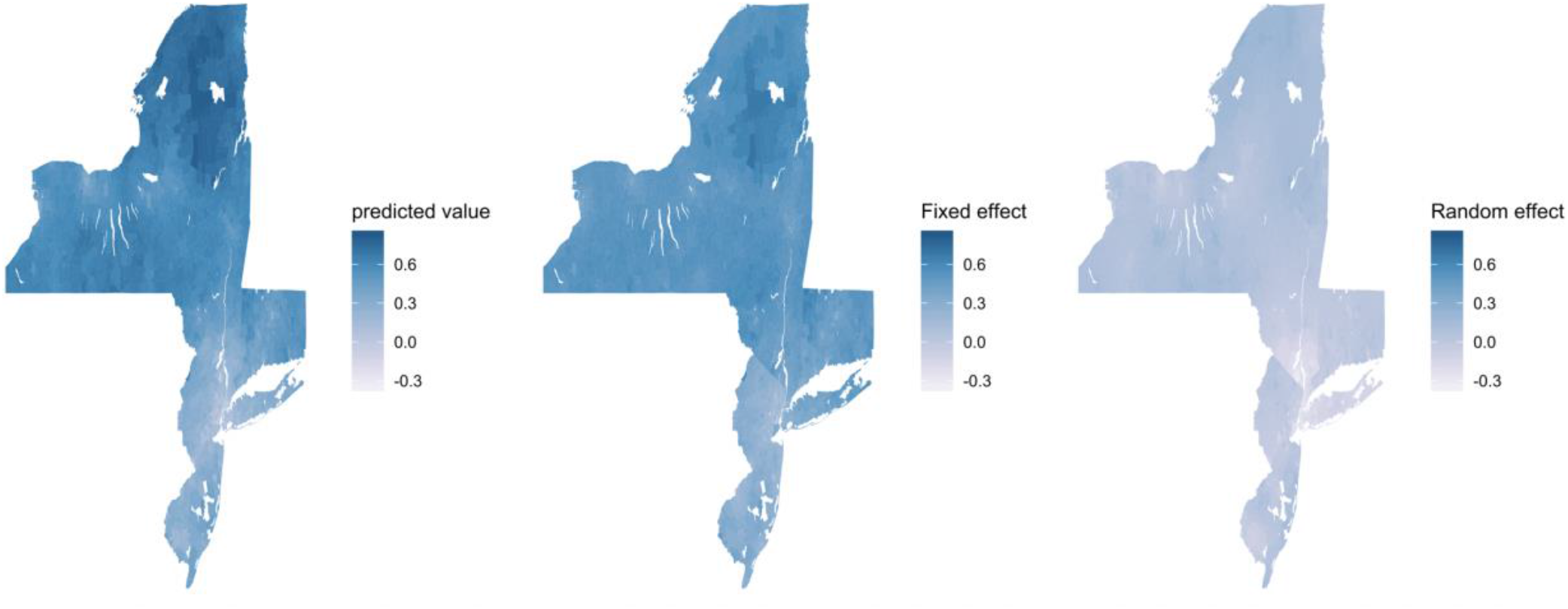
Maps comparison among 2_nd_ stage predicted value, fitted value based on fixed effect, and residuals explained by random effect.

**Supplement Table 2.** Percent of residual variation attributable to each random effect in RSV epidemics in New Jersey, New York, and Connecticut in the best fit model

|  | New Jersey | New York | Connecticut |
| --- | --- | --- | --- |
| School District | 7% (3%, 14%) | 6% (3%, 12%) | 32% (13%, 58%) |
| Spatial Effects | 93% (86%, 97%) | 94% (88%, 97%) | 68% (42%, 87%) |

**Supplement Figure 5.**
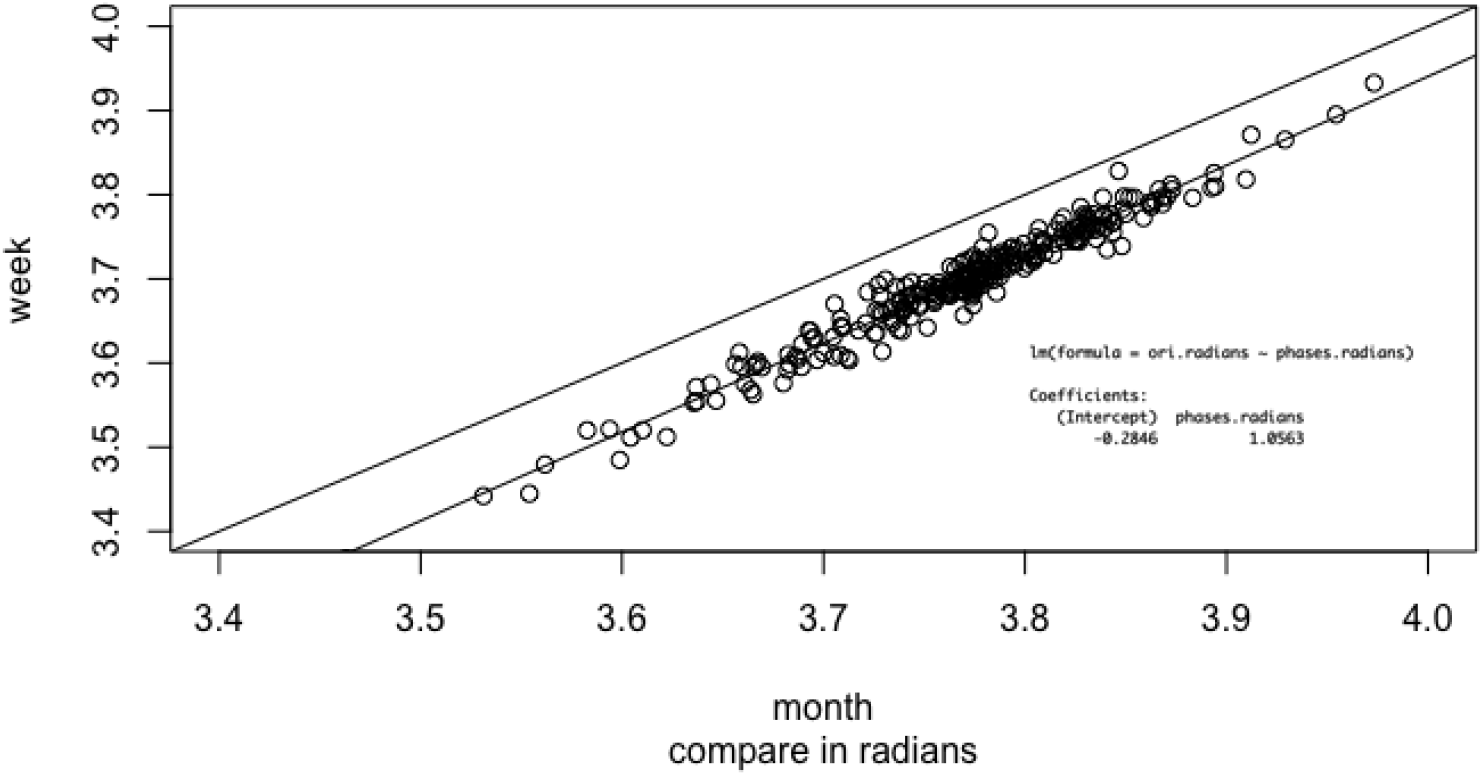
Posterior estimation using weekly data vs monthly data in Connecticut. The regression coefficient 1.06 showed that phase estimates using aggregated monthly data is consistent with using weekly data.

